# Multi-sectoral Coordination During the COVID-19 Pandemic: Practices, Challenges, and Recommendations for Future Preparedness - A Systematic Literature Review Protocol

**DOI:** 10.1101/2025.01.08.25320177

**Authors:** Javan Solomon Okello, Themba Ginindza, Julius Nyerere

## Abstract

**Introduction:** the COVID-19 pandemic amplified the need for robust multi-sectoral coordination; yet the specific mechanisms, benefits, and challenges of such collaboration particularly in low-and middle-income countries (LMICs) remain poorly synthesised.

**Objective:** To identify the key elements, benefits, challenges, and improvement strategies of multi-sectoral coordination during COVID-19, with comparative insight between LMICs and high-income countries (HICs).

**Eligibility criteria:** Empirical studies (qualitative, quantitative, or mixed-methods) published in English between 1 January 2020 and 15 August 2024 that examine any coordination mechanism (e.g., task forces, public-private partnerships, inter-agency committees) related to COVID-19 response.

**Information sources:** PubMed, EBSCOhost, Emerald Insight, Google Scholar, and targeted grey-literature repositories will be searched; reference lists and citation chaining will supplement database queries.

**Risk-of-bias assessment:** Two reviewers will independently appraise qualitative studies with CASP and non-randomised quantitative studies with ROBINS-I; disagreements will be resolved by consensus or third-reviewer adjudication.

**Data synthesis:** Owing to the anticipated dominance of qualitative evidence, a CFIR-anchored framework synthesis will be conducted. Quantitative findings will be narratively summarised and, where outcomes are commensurate, explored for fixed-effect pooling. Subgroup analyses will contrast LMIC versus HIC contexts; sensitivity analyses will exclude studies at serious/critical risk of bias. Confidence in cumulative evidence will be graded with CERQual (qualitative) and GRADE (quantitative).

**Ethics and dissemination:** No new human data will be collected; therefore additional REC approval is unnecessary. The overarching PhD project holds approvals from the University of KwaZulu-Natal (BREC/00007520/2024) and Kenya’s NACOSTI (NACOSTI/P/24/37716). Results will be disseminated via open-access publication, conference presentation, and policy briefs to Nairobi County health stakeholders.

**Strengths and Limitations of This Study:**

- Comprehensive multi-database and gray-literature searching will minimise retrieval bias
- Two reviewers will independently screen, extract and appraise studies, enhancing methodological rigour.
- Framework synthesis mapped to the consolidated framework for implementation research (CFIR) domains provides a transparent, theory-driven structure for integrating heterogeneous qualitative evidence.
- CASP and ROBINS-I instruments will standardise risk-of-bias assessment across qualitative and non-randomised quantitative studies.
- **Limitation:** English-language restriction and the paucity of quantitative studies preclude meta-analysis and may introduce language bias.

## 2. Introduction

### 2.1 Rationale

The COVID-19 pandemic exposed the fragility of global health systems and highlighted the critical role of multi-sectoral coordination in addressing complex public health crises. Effective coordination between stakeholders such as governments, health systems, private enterprises, civil society, and academia has been pivotal in managing the pandemic’s far-reaching impacts (1). However, despite its recognized importance, multi-sectoral coordination remains inadequately understood, particularly in low-and middle-income countries (LMICs), where systemic and structural barriers often hinder collaboration (2).

During the COVID-19 pandemic, intergovernmental collaboration, global partnerships, decentralized humanitarian efforts, digital knowledge-sharing platforms, task forces, and public-private partnerships were used as mechanisms for multisectoral coordination (3–6). Existing research points to key gaps in understanding how these mechanisms drive effective coordination during pandemics. Fragmented governance, siloed decision-making, and inconsistent communication have been cited as persistent challenges in LMICs, exacerbating the difficulties of resource mobilization, equitable service delivery, and policy implementation (7,8). While high-income countries (HICs) have benefited from robust emergency systems and centralized governance frameworks, LMICs such as Kenya face fragmented authority and limited digital infrastructure, which restrict the effectiveness of coordination efforts (9,10). Moreover, urban LMIC settings like Nairobi County present unique challenges, including high population density, resource inequities, and socio-economic diversity, which require tailored approaches to multi-sectoral collaboration.

This systematic review is necessary to address these critical knowledge gaps. By synthesizing evidence on the mechanisms, benefits, and challenges of multi-sectoral coordination during the COVID-19 pandemic, the study aims to provide actionable recommendations to strengthen future pandemic preparedness. The findings will contribute to global efforts to build resilient public health systems while offering context-specific insights for LMICs where the need for effective coordination is most urgent. Addressing these gaps will not only enhance pandemic preparedness but also inform strategies for tackling other complex public health challenges in resource-constrained settings.

### 2.2 Objectives

The primary objective of this systematic review is to comprehensively examine the key elements, benefits, and challenges of multi-sectoral coordination during the COVID-19 pandemic. Additionally, the review aims to identify and evaluate strategies to enhance coordination mechanisms for future public health emergencies. To achieve these objectives, a hybrid framework combining the PICO and SPIDER methodologies will be employed, facilitating the integration of both quantitative and qualitative evidence.

#### Research Questions Frameworks

##### PICO Framework (Quantitative Focus)

- **Participants:** Stakeholders involved in multi-sectoral coordination, including government officials, health agencies, private sector actors, civil society organizations, and academic institutions.
- **Interventions:** Coordination mechanisms such as task forces, public-private partnerships, incident management systems, and inter-agency committees.
- **Comparators:** Contextual comparisons between Low-and Middle-Income Countries (LMICs) and High-Income Countries (HICs) to identify differences in coordination mechanisms and outcomes.
- **Outcomes:** Evaluation of the key elements of coordination mechanisms, observed benefits (e.g., improved resource mobilization, enhanced governance), challenges (e.g., fragmented governance, communication breakdowns), and actionable recommendations for strengthening multi-sectoral coordination.

##### SPIDER Framework (Qualitative and Mixed-Methods Focus)

- **Sample:** Include stakeholders from both LMICs and HICs involved in multi-sectoral coordination during the COVID-19 pandemic. This includes government officials, health agencies, private sector actors, civil society organizations, and academia in each context.
- **Phenomenon of Interest:** Multi-sectoral coordination during the COVID-19 pandemic, with an emphasis on how these processes, mechanisms, and factors differ or align between LMICs and HICs.
- **Design:** Include studies that explore multi-sectoral coordination through qualitative methodologies (e.g., interviews, case studies) and mixed-methods approaches, ensuring that these designs facilitate comparisons between LMIC and HIC contexts.
- **Evaluation:** Assess and compare the key elements, benefits, and challenges of multi-sectoral coordination in LMICs versus HICs. This includes evaluating the effectiveness of different strategies and contextual factors that influence these outcomes.
- **Research Type:** Include both descriptive studies that outline the elements and challenges of coordination in each context and explanatory studies that explore the underlying reasons for differences and similarities between LMICs and HICs.

## Research Questions

### 1. Overarching question

What are the key elements, benefits, challenges, and improvement strategies associated with multi-sectoral coordination during the COVID-19 pandemic?

#### Operational sub-questions

i. **RQ 1** How did specific coordination mechanisms (task forces, PPPs, incident-management systems) influence resource-mobilisation and governance effectiveness in LMICs compared with HICs?
ii. **RQ 2** Which organisational structures, processes, and contextual factors mapped to the CFIR domains facilitated or impeded effective coordination?
iii. **RQ 3** What actionable strategies have stakeholders proposed or implemented to strengthen future multi-sectoral pandemic preparedness, and what evidence exists for their feasibility?

#### Expected Contributions

Addressing these research questions through a CFIR framework will enable the review to bridge critical gaps in understanding and practice related to multi-sectoral coordination in public health crises. The findings will offer a robust synthesis of existing evidence, serving as a valuable resource for policymakers, researchers, and practitioners. This comprehensive analysis will inform efforts to enhance multi-sectoral collaboration, thereby improving responses to future public health emergencies

## 3. Methods

### 3.1 Eligibility Criteria

Guided by both the PICO (Population, Intervention, Comparator, Outcome) and SPIDER (Sample, Phenomenon of Interest, Design, Evaluation, Research Type) frameworks, this systematic review will include studies that investigate multi-sectoral coordination during the COVID-19 pandemic and meet rigorous inclusion and exclusion benchmarks. Specifically, eligible studies must:

1. **Focus on Multi-Sectoral Coordination Mechanisms**: Address at least one of the following core themes: coordination elements, benefits, or challenges facing interventions such as government-led task forces, public–private partnerships, or inter agency collaborations (PICO: Intervention/ SPIDER: Phenomenon).
2. **Target Relevant Populations/Samples**: Involve stakeholders such as government officials, private sector entities, civil society organizations, or academic institutions (PICO: Population/ SPIDER: Sample).
3. **Provide Empirical Evidence**: Present primary data or robust secondary analyses that offer quantitative, qualitative, or mixed-methods insights (SPIDER: Design, Research Type).
4. **Outcomes/Evaluation**: Include outcomes that illuminate the effectiveness, barriers, or enabling factors of coordination, offering recommendations for strengthening future pandemic responses (PICO: Outcomes/ SPIDER: Evaluation).
5. **Publication Type and Quality**: Be peer-reviewed journal articles or credible gray literature (e.g., government/WHO reports) published in English from the start of COVID-19 in 2020 through August 2024.

Exclusion criteria comprise editorials, commentaries, opinion pieces, and materials unrelated to multi-sectoral coordination during COVID-19.

### 3.2 Information Sources

To ensure comprehensive coverage, the review will search multiple electronic databases and gray literature sources. Databases to be included are EBSCO Host (CINAHL, MEDLINE,

PsycINFO), Emerald Insight, PubMed, and Google Scholar. Gray literature will include reports and publications from reputable organizations such as the WHO and government bodies. The search will focus on publications dating from 1^st^ January 2020 to 15^th^ August 2024, reflecting the evolution of multi-sectoral coordination during the pandemic.

### 3.3 Search Strategy

The lead researcher and the co-authors designed the database strategies in line with the PRISMA-S reporting checklist. Four bibliographic sources will be searched from 1 January 2020 to 15 August 2024: PubMed/MEDLINE, EBSCOhost (Global Health + CINAHL), Emerald Insight, and Google Scholar. The PubMed strategy combines controlled vocabulary (MeSH) and free-text terms for (i) *multi-sectoral coordination*, (ii) *evaluation constructs*, and (iii) *COVID-19*. Language is limited to English. No study-design filters are applied. Search strategies for the other three databases were translated using the PRESS checklist to preserve concepts and syntax. For Google Scholar, only the first ten result pages (≈ 100 records) will be screened to balance sensitivity and feasibility.

The full line-by-line strategies, with search dates, record yields, and query translations for each database, are provided in Supplementary Table S1 to permit complete reproducibility.

### 3.4 Data-collection process

All citations retrieved from the finalized search strategy will be exported to EndNote 20, which will serve as the master reference library and the single source of truth for de-duplication. The de-duplicated records will be uploaded into Covidence for title/abstract screening and subsequent full-text appraisal. Two reviewers will screen each record independently; disagreements will be adjudicated by a third reviewer. Prior to full rollout, a pilot exercise involving 5 % of the search yield will test the eligibility criteria, and inter-rater reliability will be computed (Cohen’s κ). A threshold of κ ≥ 0.70 has been set as the minimum acceptable concordance; wording ambiguities in the criteria will be refined until this standard is achieved. All full-text PDFs will be stored on an access-controlled Google Drive directory, with automatic version history retained for audit purposes.

Data extraction will proceed in a two-tier workflow. First, customised Python scripts and an LLM-enabled PDF parser will pre-populate a structured Excel/Google Sheets template with bibliographic details and predefined data items (sectoral involvement, coordination level, mechanism type, inclusivity/transparency attributes, resource-allocation modalities, quantified benefits, qualitative outcomes, challenges, mitigation strategies, practice-oriented recommendations). Two reviewers will then independently verify and complete the template, with a second reviewer cross-checking all critical variables; unresolved discrepancies will be resolved by a third reviewer. Verbatim qualitative excerpts pertaining to coordination determinants will be imported into NVivo 14 and coded deductively against the five CFIR domains, with inductive codes added when necessary. Risk-of-bias appraisal will be conducted in parallel: CASP for qualitative reports and ROBINS-I for the two non-randomised quantitative studies, after a calibration exercise to harmonise scoring. Authors of primary studies will be contacted via e-mail for missing or unclear information, with a 21-day response window; non-responses will be logged as “unavailable.”

All live extraction files, appraisal forms, and amendment logs will reside on an encrypted local drive mirrored to the Google Drive repository. Access is restricted to the lead investigator and two doctoral supervisors, and an automated weekly back-up routine ensures data integrity and disaster recovery.

### 3.5 Data Items

This review will extract and analyze variables central to understanding the contextual and operational features of multi-sectoral coordination during the COVID-19 pandemic. Key data items include the sectors involved (e.g., health, education, private sector), levels of coordination (e.g., local, national, international), and coordination mechanisms such as task forces and public-private partnerships. The review will examine the components and features of these mechanisms, with a focus on attributes like inclusivity, transparency, and timeliness in decision-making processes. Other data items include resource allocation methods, emphasizing how human, financial, and logistical resources were managed. The benefits of multi-sectoral coordination will be captured through both quantitative outcomes, such as improved efficiency in pandemic responses, and qualitative outcomes, such as strengthened governance frameworks. Challenges and barriers to coordination, will also be identified alongside strategies employed to mitigate these issues. Finally, recommendations for both practice and research will be included, providing actionable insights for future improvements.

### 3.6 Outcomes and Prioritization

The primary outcomes of interest for this review are the identification of key elements and mechanisms of multi-sectoral coordination, and their contributions to improved public health responses, equitable resource allocation, and effective governance during the COVID-19 pandemic. Specific focus will be placed on outcomes such as enhanced efficiency in pandemic response mechanisms, strengthened decision-making processes, and the mitigation of barriers like fragmented authority and communication breakdowns. These outcomes will inform actionable recommendations for improving coordination frameworks, particularly in LMICs like Kenya.

Secondary outcomes will explore the long-term benefits of multi-sectoral coordination, including the institutionalization of coordination mechanisms for future pandemic preparedness and enhanced trust in governance systems. Additional insights will be drawn from cross-sectoral synergies, such as the role of education systems in supporting health responses and civil society in addressing inequities. Variations in outcomes across local, national, and international coordination levels will also be examined, providing a nuanced understanding of how contextual factors and governance structures influence the effectiveness of multi-sectoral approaches.

### 3.7 Risk of Bias Assessment

Two reviewers will appraise each study independently, first calibrating on a pilot set of five papers to harmonise interpretation of signalling questions. Disagreements will be resolved by consensus or, if required, a third-reviewer adjudication.

#### Qualitative evidence: CASP

The Critical Appraisal Skills Programme (CASP) Qualitative Checklist (10 items) will be used. For each item the response options are *Yes / No / Unclear*.

- **Domain rating:** An item answered *Yes* is coded “low concern,” *No* “high concern,” and *Unclear* “some concern.”
- Overall study rating:

`o` **Low risk:** ≥ 8 of 10 items “low concern” **and** none “high concern.”
`o` **Moderate risk:** 5–7 items “low concern” **or** exactly one “high concern.”
`o` **High risk:** ≤ 4 items “low concern” **or** ≥ 2 items “high concern.”

#### Quantitative evidence: ROBINS-I

The Risk Of Bias In Non-randomised Studies of Interventions (ROBINS-I) tool will be applied to the non-randomised quantitative studies across its seven domains. Each domain is rated Low, Moderate, Serious, Critical, or No information following Cochrane Handbook guidance.

- **Overall study rating:** The worst domain rating determines the overall assessment (e.g., one “Serious” domain will be interpreted as overall “Serious”).
- Where outcome-specific bias differs from study-level bias, an additional outcome-level tag (e.g., “Serious-Outcome”) will be noted in the extraction sheet.

#### Use of bias ratings in synthesis

- Studies at High/Critical risk will be excluded from any exploratory pooling and flagged during framework synthesis.
- Sensitivity analyses will be run with and without High/Critical studies to test robustness.
- Risk-of-bias judgements will directly inform CERQual (qualitative) and GRADE (quantitative) certainty assessments.

All appraisal forms, piloting notes, and adjudication logs will be stored with date-stamped version history on the encrypted project drive and made available as supplementary material upon publication.

### 3.8 Data Synthesis

#### Qualitative synthesis approach

The study will undertake a framework synthesis guided by the Consolidated Framework for Implementation Research (CFIR). After familiarisation with the extracted material, an a-priori analytical framework comprising the five CFIR domains such as Intervention Characteristics, Outer Setting, Inner Setting, Characteristics of Individuals, and Process will be used to index all qualitative findings. Two reviewers will independently allocate verbatim excerpts to relevant domains and sub-constructs, recording justifications in NVivo 14. Discrepancies will be resolved through discussion or third-reviewer arbitration. Once indexing is complete, data will be charted into a domain-by-study matrix; emergent sub-themes that do not fit the original framework will be inductively added and clearly marked. Finally, the study will map and interpret patterns within and across domains, paying special attention to contextual contrasts between LMIC and HIC settings and to the coherence with the limited quantitative evidence

### Quantitative synthesis approach

Where quantitative data are available, results will first be tabulated to show the study context, sample, coordination mechanism, outcome metric, effect estimate, and risk-of-bias rating (ROBINS-I).

**Criteria for pooling.** A meta-analysis will be undertaken only if two or more studies report an identical outcome in comparable units, and each study is rated low or moderate overall risk of bias. Should three or more studies meet criteria, heterogeneity will be assessed with the χ² test and quantified with **I²** (low < 30 %, moderate 30–60 %, substantial > 60 %). If I² > 60 %, a random-effects (DerSimonian–Laird) model will be applied; otherwise, fixed-effect pooling will be retained. Analyses will be run in R (meta v6.5-0); results will be displayed via forest plots. If ten or more studies contribute to a meta-analysis, small-study effects will be explored with funnel plots and Egger’s test. Subgroup and sensitivity analyses described above will be repeated in the meta-analytic framework to probe heterogeneity drivers.

**CFIR integration.** Each quantitative outcome will be mapped to the most relevant CFIR domain, permitting cross-walk with the qualitative matrix and facilitating convergent interpretation.

### Subgroup and sensitivity analyses

To illuminate contextual contingencies, a subgroup comparison will contrast effect directions between low-and middle-income countries (LMICs) and high-income countries (HICs), explicitly noting how CFIR domains manifest differently across resource settings. Sensitivity analyses will (i) exclude studies at serious or critical risk of bias, (ii) re-run descriptive syntheses after omitting studies with outlying sample sizes or non-comparable outcome definitions, and

(iii) test the influence of analytic choices such as fixed-effect versus narrative reporting. These steps will enhance the robustness and contextual relevance of the quantitative strand, while ensuring coherence with the review’s primary objective of elucidating the elements, benefits, challenges, and improvement strategies for multi-sectoral coordination during the COVID-19 pandemic.

### 3.9 Meta-Bias Assessment

#### 3.9.1 Publication bias

**Quantitative evidence.** If ten or more studies report on the same outcome, we will assess publication bias using two methods. First, funnel plots will be visually inspected for asymmetry. Second, Egger’s regression test (two-tailed, p < 0.10) will be conducted using the metabias function in R. Any observed asymmetry will be interpreted cautiously, taking into account study sample sizes and between-study heterogeneity before attributing it to publication bias.

**Qualitative evidence.** The search strategy already incorporates grey literature and pre-print servers; nevertheless, we will document any domain where only peer-reviewed sources surface and discuss the implication for transferability.

#### 3.9.2 Selective-reporting bias

For each included study the outcomes listed in Methods will be cross-checked against those reported in Results. Discrepancies will be logged in an *Outcome-Level Bias* column of the extraction sheet, using an adapted ORBIT decision tree. Evidence of suppression or outcome switching will feed into the risk-of-bias judgement and downgrade certainty (see Section 3.9.4).

#### 3.9.3 Missing-data protocol

For any critical missing information, we will first email the corresponding authors, sending one follow-up reminder and then consider it a non-response if the author does not get back to us after 21 days. In quantitative studies, if dispersion measures are unavailable but means and sample sizes are reported, standard deviations will be imputed with established formulas; studies whose effect sizes remain unreconstructable will be retained only in the narrative synthesis. In qualitative studies, absent data will be flagged as “data absent” within NVivo, and this deficiency will be accounted for in the adequacy domain of the CERQual assessment. Every action and decision will be documented in the Amendment Log, and sensitivity analyses will exclude studies that rely on imputed or unrecoverable data.

#### 3.9.4 Confidence in Cumulative Evidence Quantitative findings: GRADE

For each pooled or narratively summarised outcome we will rate the certainty as High, Moderate, Low, or Very Low by sequentially considering: (i) risk of bias (ROBINS-I), (ii) inconsistency (I² or descriptive divergence), (iii) indirectness, (iv) imprecision (95 % CI width, optimal-information size), and (v) publication bias. Observational studies will start at “Low” and be upgraded for large effect, dose–response, or residual-confounding plausibility. A “Summary of Findings” table will be generated in GRADEpro GDT.

##### Qualitative findings: CERQual

Each thematic finding emerging from the CFIR-based framework synthesis will be assessed across four domains: methodological limitations, coherence, adequacy, and relevance yielding a confidence rating of High, Moderate, Low, or Very Low. Explanatory footnotes will justify any downgrades. A CERQual evidence profile will accompany the results.

### 3.10 Patient and public involvement

Patients and members of the public were not involved in the design, conduct, reporting or dissemination plans of this systematic-review protocol.

### 4.0 Amendments

According to this study an amendment is defined as any post-registration change that substantively alters the eligibility criteria, search strategy, analytic plan, outcomes, or authorship configuration; purely typographical or formatting corrections are treated as administrative edits. All substantive modifications will be processed through a four-step pathway. First, the lead investigator will circulate the proposed change to the full author team for a 72-hour commentary period; unresolved disagreements will be escalated to a third-party adjudicator. Second, once consensus is reached, the master protocol maintained on medRxiv will be updated and re-tagged sequentially (v1.1, v1.2, etc.), thereby generating a date-stamped DOI for each iteration. Third, the details will be entered into a stand-alone Amendment Log that records the date, protocol section affected, original and revised text, rationale, approving initials, and anticipated impact on the analysis; the Monitoring-and-Evaluation officer cross-checks the log monthly for completeness. Fourth, any amendment that changes eligibility criteria, outcomes, or the analytic plan will be mirrored in the PROSPERO record within seven days, explicitly citing the relevant medRxiv version.

Public transparency is ensured in two ways: (i) the final manuscript will include a concise narrative summary of all substantive amendments with hyperlinks to the medRxiv version history, and (ii) the full Amendment Log will be provided as supplementary material. A quarterly internal audit will verify concordance between the Log, medRxiv tags, and PROSPERO history, with findings documented in a one-page quality-assurance memo. Should an amendment arise after data extraction has begun, its potential to introduce bias will be flagged in the Log’s decision-impact field and, if non-trivial, discussed in the Limitations section of the final review. In emergencies such as database deprecation the 72-hour consultation window may be bypassed, but retrospective ratification must occur within one week.

## 5. Ethics and Dissemination

### Ethics Approval

This systematic literature review synthesises data already in the public domain; consequently, no additional research-ethics committee (REC) approval is required for this component. The review constitutes Objective 1 of a broader PhD project that will subsequently collect primary qualitative and quantitative data in Nairobi County. For that future fieldwork, overarching approvals have been obtained from the University of KwaZulu-Natal Biomedical Research Ethics Committee (BREC, Ref. BREC/00007520/2024, 25 Nov 2024) and Kenya’s National Commission for Science, Technology & Innovation (NACOSTI, Licence NACOSTI/P/24/37716). These approvals are noted here for transparency but were not prerequisites for the present review.

### Dissemination Plan

#### Target Audience

The protocol will be disseminated to key stakeholders in public health, governance, and research to foster engagement and collaboration. Primary audiences include researchers, academics, and policymakers involved in pandemic preparedness and multi-sectoral coordination. Special emphasis will be placed on reaching stakeholders in LMICs, including Nairobi County officials, public health practitioners, and regional bodies such as the African Union. International organizations such as WHO and other global health actors will also be targeted to encourage alignment with broader pandemic preparedness strategies.

#### Publication

The protocol will be submitted to a high-impact, peer-reviewed journal specializing in public health or global health research, such as *BMJ Open*. Open-access publication will be prioritized to ensure that the protocol is widely accessible to researchers, practitioners, and policymakers globally. Efforts will also be made to highlight the protocol in regional platforms, such as the *East African Medical Journal*, to ensure relevance to the Kenyan and East African contexts.

#### Conferences and Presentations

The protocol will be introduced at conferences and academic forums to engage with the global research community and promote awareness of the planned review. Opportunities will include presentations at events like the *World Congress on Public Health* and the *Global Health Systems*

*Research Symposium*. Regional conferences such as the *Kenya Health Forum* and *Africa Health Conference* will be leveraged to gather feedback and foster collaboration with local and regional stakeholders.

#### Non-Academic Channels

The protocol will be disseminated through institutional and organizational platforms to engage a wider audience. It will be shared on the University of KwaZulu-Natal’s website and highlighted in institutional newsletters or reports. Social media platforms such as Twitter, LinkedIn, and research-focused forums (e.g., ResearchGate) will also be used to promote the protocol and encourage engagement from the global research community.

#### Collaboration

Collaboration with key organizations such as WHO, NACOSTI, and Kenya’s Ministry of Health will be sought to amplify the reach and relevance of the protocol. Regional academic institutions and public health networks will also be engaged to encourage adoption and alignment with ongoing pandemic response research.

#### Timing

The protocol will be disseminated immediately upon acceptance and publication in a peer-reviewed journal. Subsequent presentations and engagements will align with key global and regional events, ensuring maximum visibility and relevance.

## Data Availability

All data produced in the present study are available upon reasonable request to the authors

## 6. Acknowledgments

I would like to express my deepest gratitude to my primary supervisor, Prof. Themba, and co-supervisor, Dr. Julius, for their unwavering guidance, constructive feedback, and mentorship throughout the development of this systematic review protocol. I extend my heartfelt thanks to Dr. Bonface Oyugi of WHO AFRO for his invaluable insights into systematic literature reviews and for providing access to critical resources on multi-sectoral coordination. I am also deeply indebted to Dr. Yasushi Sawazaki, whose mentorship and inspiration shaped my academic pursuits, and to my wife, Dr. Vivian Nyaata of Kisii University, for her steadfast support, encouragement, and thoughtful insights.

This work would not have been possible without the institutional support provided by the University of KwaZulu-Natal (UKZN), particularly the Department of Public Health, whose resources and infrastructure facilitated this research. I also wish to acknowledge the ethical oversight and contributions of Prof. Karama of AMREF Kenya, whose guidance greatly streamlined the process of obtaining clearance from NACOSTI.

A special thanks is extended to my colleagues, family, and friends who offered moral support during this endeavor. Lastly, I dedicate this protocol to my boss, Dr. Yasushi Sawazaki, whose leadership during the JICA-funded East African Community Regional COVID-19 Countermeasures at the Border Posts research project provided me with the foundational insights and inspiration to focus on multi-sectoral coordination in pandemic response as my PhD research topic. His mentorship has been instrumental in shaping my understanding of the critical role of collaboration in addressing public health crises.

## 7 Conflict of Interest

The authors declares no conflict of interest in relation to the development of this systematic review protocol. The research was conducted independently, and no external parties influenced the study design, methods, or content.

Institutional support was provided by the University of KwaZulu-Natal, specifically the Department of Public Health, to facilitate the research. However, this support did not exert any undue influence on the study’s objectives, methodology, or anticipated outcomes.

Ethics approvals obtained from the University of KwaZulu-Natal Biomedical Research Ethics Committee (BREC) and Kenya’s National Commission for Science, Technology & Innovation (NACOSTI) were carried out in compliance with independent regulatory standards, ensuring the integrity of the research process.

All collaborators, including mentors and supervisors, provided guidance strictly within their academic and professional capacities, without any competing interests affecting the development or direction of this protocol.

## 8. Author Statement

This systematic review protocol was developed through the collaborative efforts of all listed authors, with each contributing significantly to the study. Javan Solomon Okello conceptualized the review, developed the methodology, conducted the initial drafting of the protocol, and ensured its alignment with the study objectives. Prof. Themba and Dr. Julius provided supervision and guidance throughout the development process, critically reviewing the protocol and offering intellectual input to enhance its rigor and coherence. Dr. Bonface Oyugi of WHO AFRO contributed expert insights into systematic literature review methodologies and facilitated access to key resources on multi-sectoral coordination.

Dr. Vivian Nyaata of Kisii University provided logistical and moral support, offering valuable feedback during the drafting and revision phases. Dr. Yasushi Sawazaki inspired the focus on multi-sectoral coordination through his mentorship and insights gained during the JICA-Funded East African Community Regional COVID-19 Countermeasures project. Prof. Karama of AMREF Kenya supported the NACOSTI clearance process by offering strategic advice and guidance.

All authors have reviewed and approved the final version of this protocol for submission and agree to be accountable for all aspects of the work, ensuring that questions related to the accuracy or integrity of any part of the work are appropriately addressed.

## 9. Data Availability Statement

This systematic review does not involve the collection of primary data. All data used in this study will be extracted from publicly available sources, including peer-reviewed journal articles and reputable gray literature. The extracted dataset, along with any technical appendices and analysis tools, will be made openly accessible through the Dryad repository. The dataset will be assigned a DOI, which will be provided upon publication. For further inquiries or access to the dataset prior to repository upload, please contact the corresponding author at

## Appendices

### Appendix A: PRISMA-P Checklist

This checklist demonstrates compliance with the PRISMA-P (Preferred Reporting Items for Systematic Review and Meta-Analysis Protocols) guidelines. Each item has been addressed in the protocol.

| SN | Section | Sub-Section | Item | Included |
| --- | --- | --- | --- | --- |
| 1 | **Administrative Information** | Title | Identify the protocol as a systematic review protocol | ☑ Yes<br>Page 1 |
|  |  | Registration | Provide the name of the registry (e.g., PROSPERO) and registration number | ☑ Yes<br>Page 1 |
|  |  | Authors | Provide names, affiliations, and contact details of all authors | ☑ Yes<br>Page 1 |
|  |  | Contributions | Specify roles of each author in protocol development | ☑ Yes<br>Page 1 |
|  |  | Support | List funding sources, sponsors, and institutional support | ☑ Yes<br>Page 1 |
|  |  | Amendments | Plan for documenting and tracking protocol amendments | ☑ Yes<br>Page 2 |
| 2 | **Introduction** | Rationale | Describe the rationale for the review in the context of existing knowledge | ☑ Yes<br>Page 6 |
|  |  | Objectives | Provide an explicit statement of the questions being addressed | ☑ Yes<br>Page 7 |
| 3 | **Methods** | Eligibility Criteria | Define inclusion and exclusion criteria (e.g., participants, interventions, outcomes, study designs) | ☑ Yes<br>Page 9 |
|  |  | Information Sources | Specify all information sources (e.g., databases, grey literature, date ranges) | ☑ Yes<br>Page 9 |
|  |  | Search Strategy | Present draft search strategy for at least one database | ☑ Yes<br>Page 10 |
|  |  | Data Management | Describe methods for managing and tracking study records | ☑ Yes<br>Page 10 |
|  |  | Study Selection Process | Outline how studies will be screened for inclusion | ☑ Yes<br>Page 10 |
|  |  | Data Collection Process | Specify data items to be extracted, including tools and processes | ☑ Yes<br>Page 10 |
|  |  | Risk of Bias Assessment | Describe tools and methods for assessing bias in included studies | ☑ Yes<br>Page 12 |
|  |  | Data Synthesis | Describe planned synthesis methods (e.g., narrative synthesis, meta-analysis) | ☑ Yes<br>Page 13 |
|  |  | Meta-Bias Assessment | Specify methods for assessing publication or reporting bias | ☑ Yes<br>Page 14 |
|  |  | Confidence in Evidence | Describe methods for assessing confidence in cumulative evidence | ☐ Yes<br>Page 14 |
|  | **Ethics and Dissemination** | Ethics Approval | Indicate whether ethics approval is required for the review | ☐ Yes<br>Page |
|  |  | Dissemination Plan | Describe plans for communicating review results (e.g., journals, conferences) | ☐ Yes<br>Page |

### Appendix B: Search Strategy

#### Research Questions

2. **Overarching question**

#### Operational sub-questions

iv. **RQ 1** How did specific coordination mechanisms (task forces, PPPs, incident-management systems) influence resource-mobilisation and governance effectiveness in LMICs compared with HICs?
v. **RQ 2** Which organisational structures, processes, and contextual factors mapped to the CFIR domains facilitated or impeded effective coordination?
vi. **RQ 3** What actionable strategies have stakeholders proposed or implemented to strengthen future multi-sectoral pandemic preparedness, and what evidence exists for their feasibility?

#### Search Strategy

The study will use a combination of primary and secondary keywords, focusing on “multisectoral coordination

((“multisectoral coordination” OR “inter-agency cooperation” OR “cross-sectoral partnership” OR “multi-stakeholder engagement” OR “collaborative governance” OR “integrated pandemic management” OR “ multi-sectoral coordination” OR “multi sectoral co-ordination” OR “multi sectoral co-ordination” OR “inter agency cooperation” OR “inter-sectoral coordination” OR “inter-sectoral collaboration” OR “collaborative coordination”) AND (“effectiveness” OR “efficacy” OR “outcomes” OR “impact” OR “success metrics” OR “performance indicators” OR “evaluation” OR “challenges” OR “barriers” OR “limitations” OR “obstacles” OR “difficulties” OR “constraints” OR “issues”)) AND (“COVID-19” OR “SARS-CoV-2” OR “coronavirus pandemic” OR “CoronaVirus” OR “severe-acute-respiratory-syndrome-related coronavirus” OR “SARS COV 2” OR “COVID 19”)

**Supplementary Table S1.** (excerpt for PubMed)

| **Line** | **Search string** | **Records (n)** |
| --- | --- | --- |
| 1 | “COVID-19”[Mesh] OR covid* OR “SARS-CoV-2” OR coronavirus* |  |
| 2 | “Intersectoral Collaboration”[Mesh] OR “multisectoral coordination” OR inter-agency cooperation OR cross-sectoral partnership* OR multi-stakeholder engagement OR collaborative governance OR taskforce* OR “public-private partnership*” |  |
| 3 | effectiveness OR efficacy OR outcome* OR impact* OR evaluation OR barrier* OR challenge* |  |
| 4 | 1 AND 2 AND 3 |  |
| 5 | Limit 4 to English and 2020:2024 |  |

**Supplementary Table S2.**
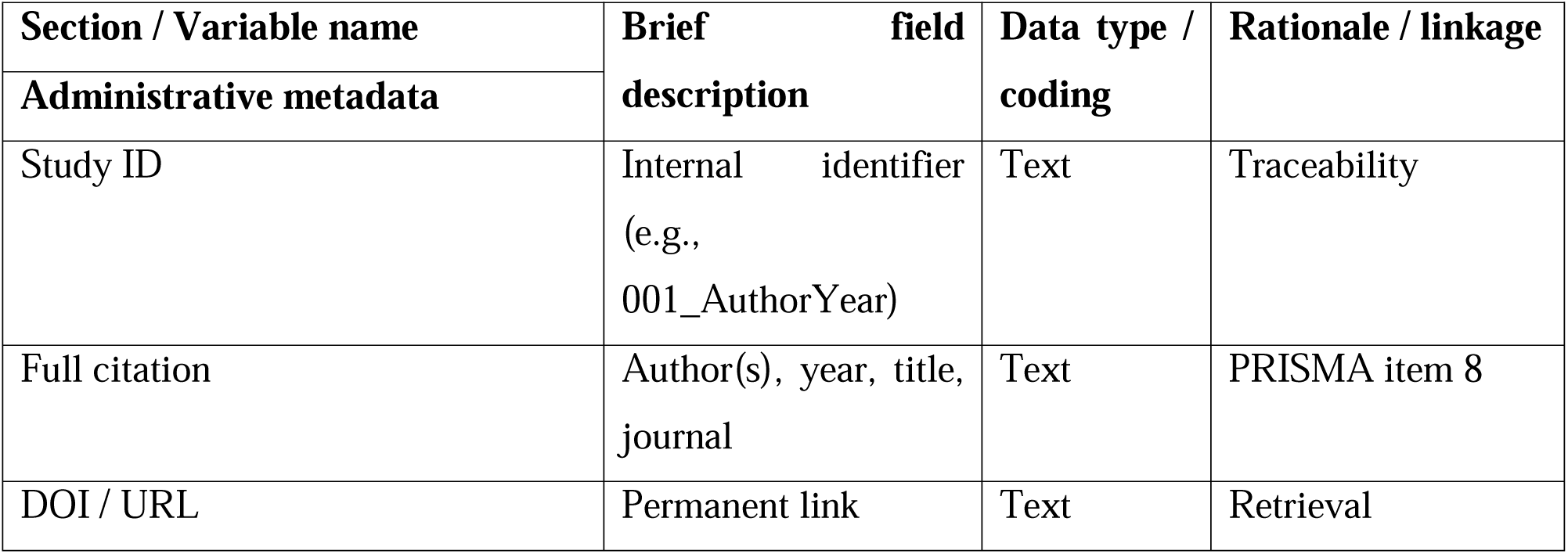
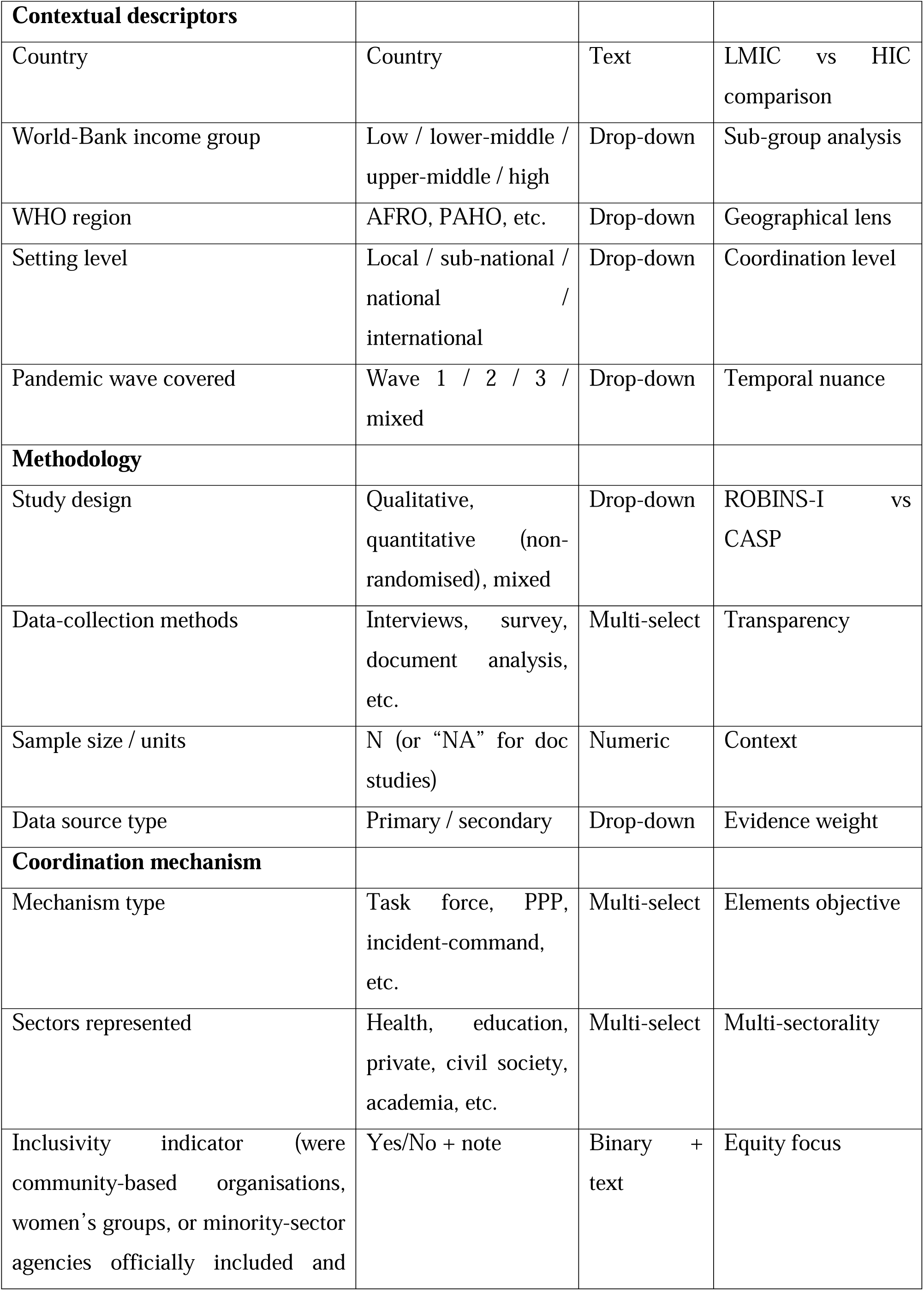
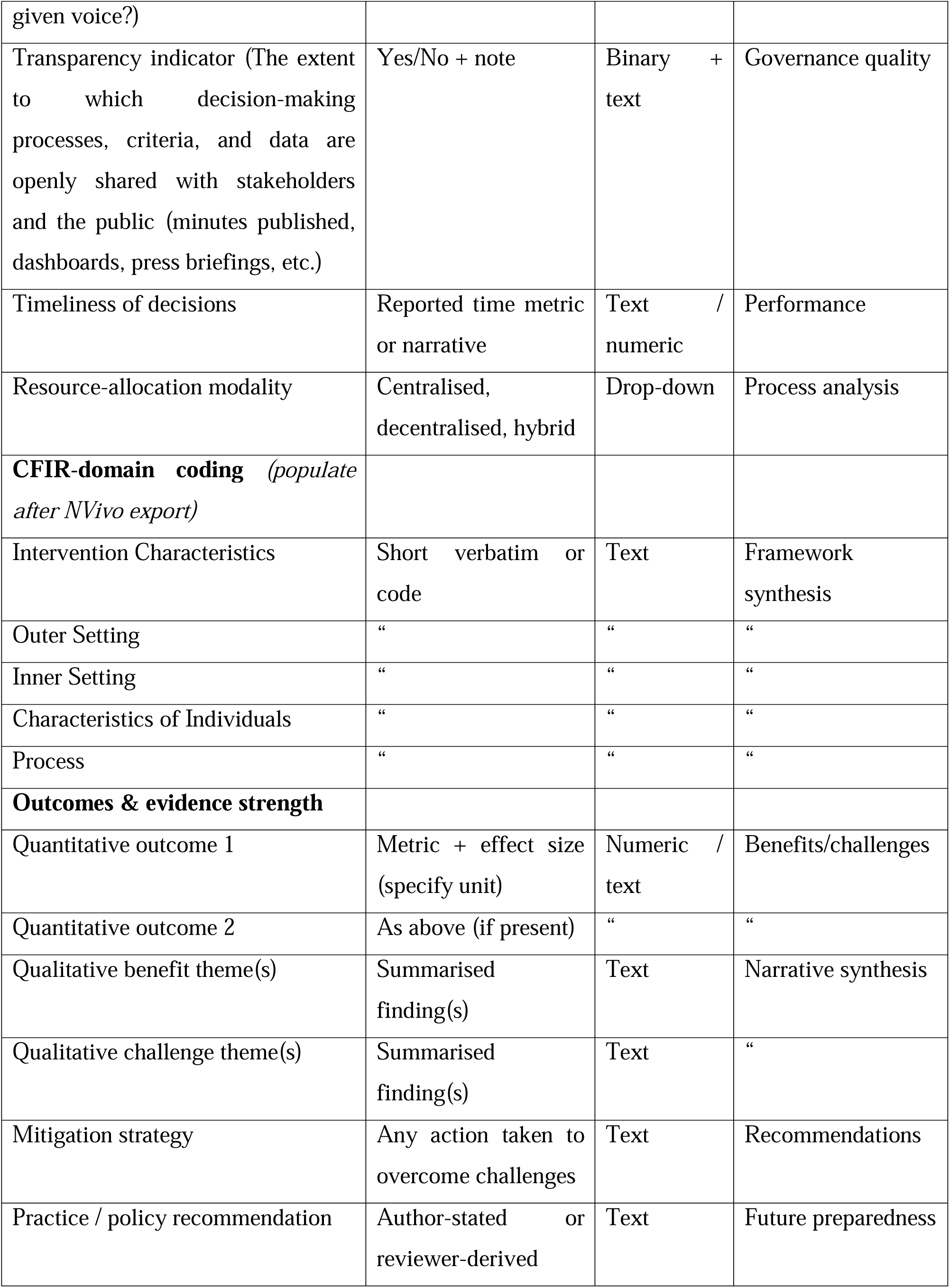
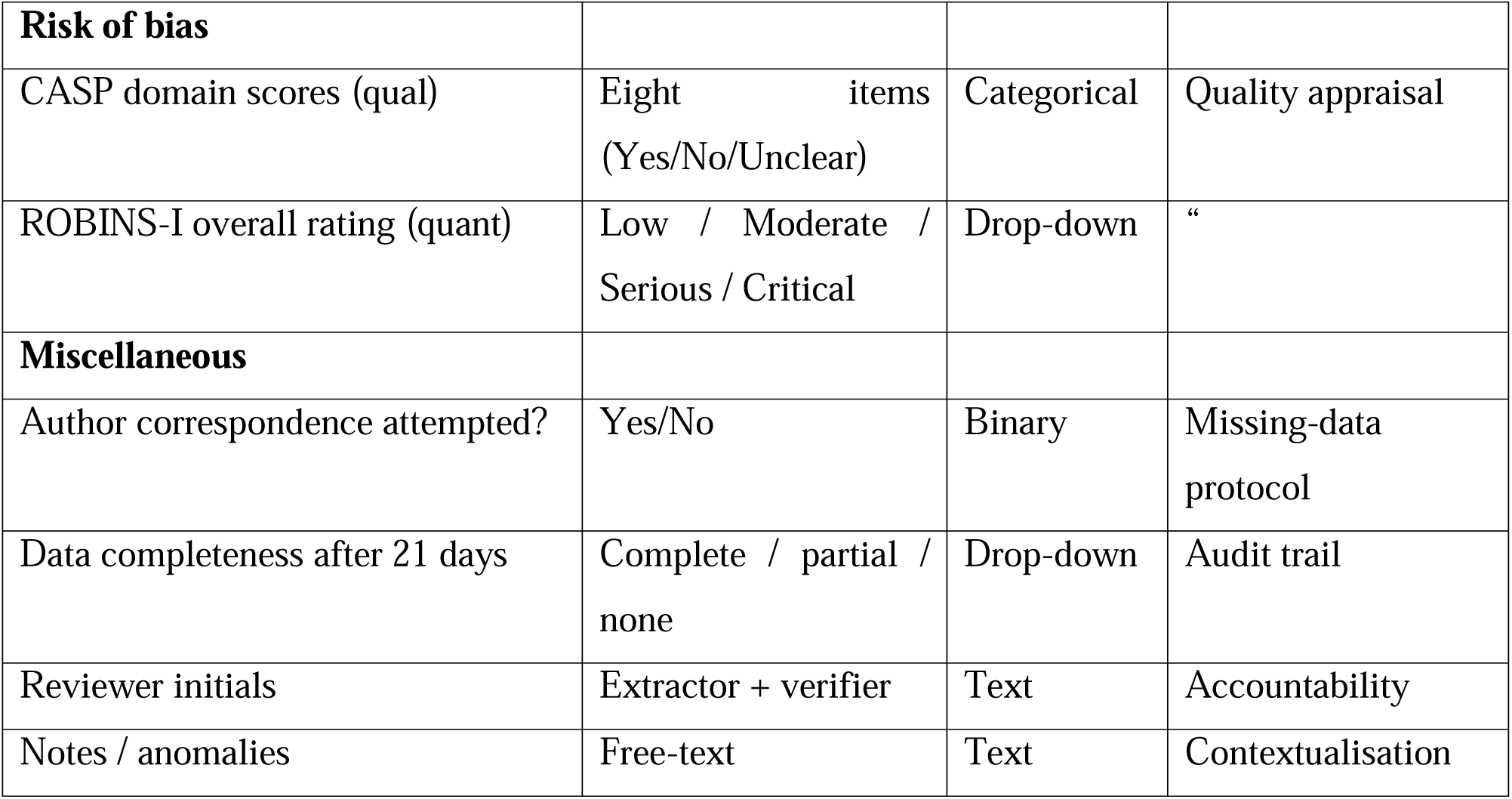
_Data-Extraction_Template.

#### Protocol amendments: documentation and tracking plan

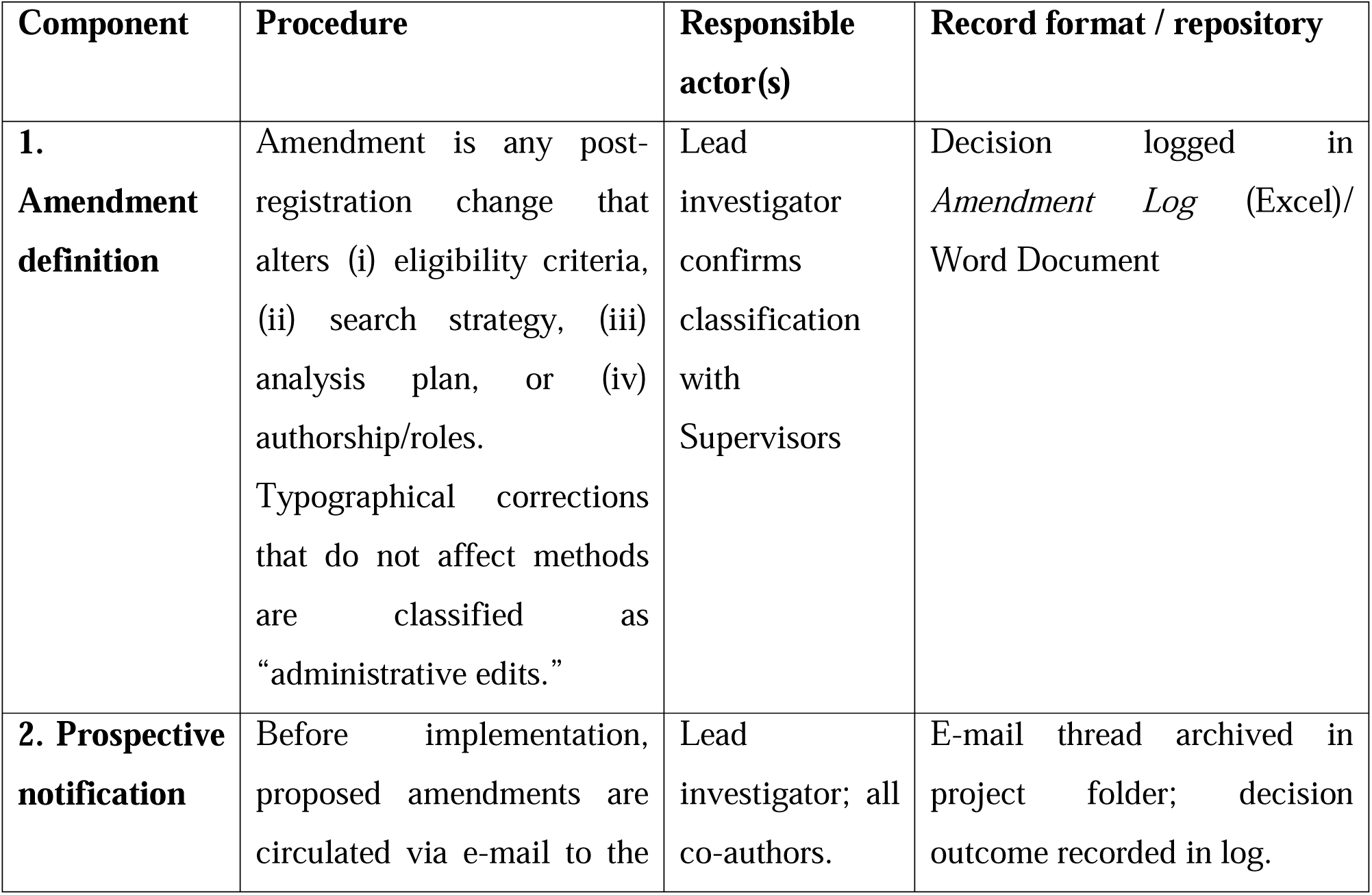
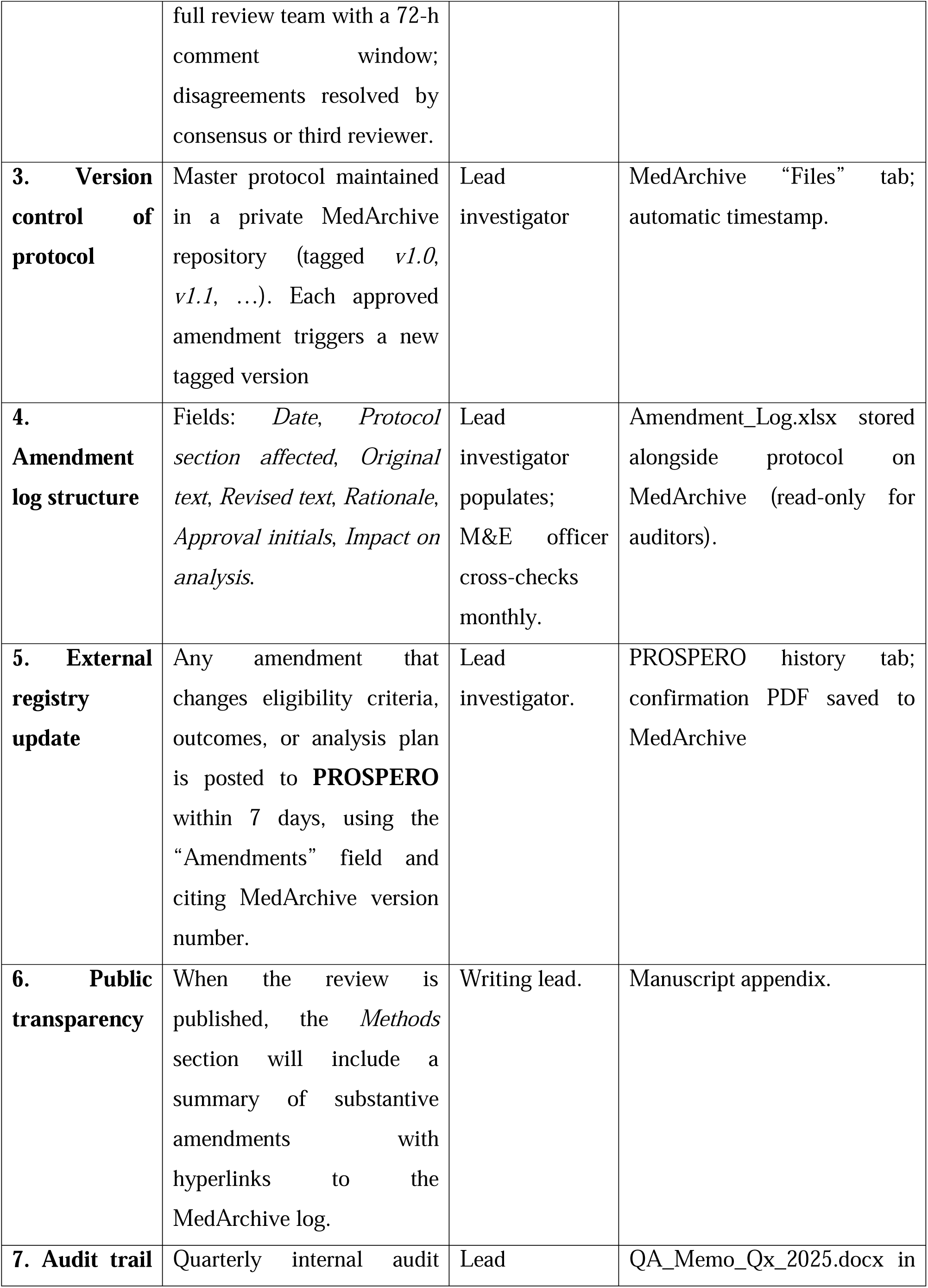
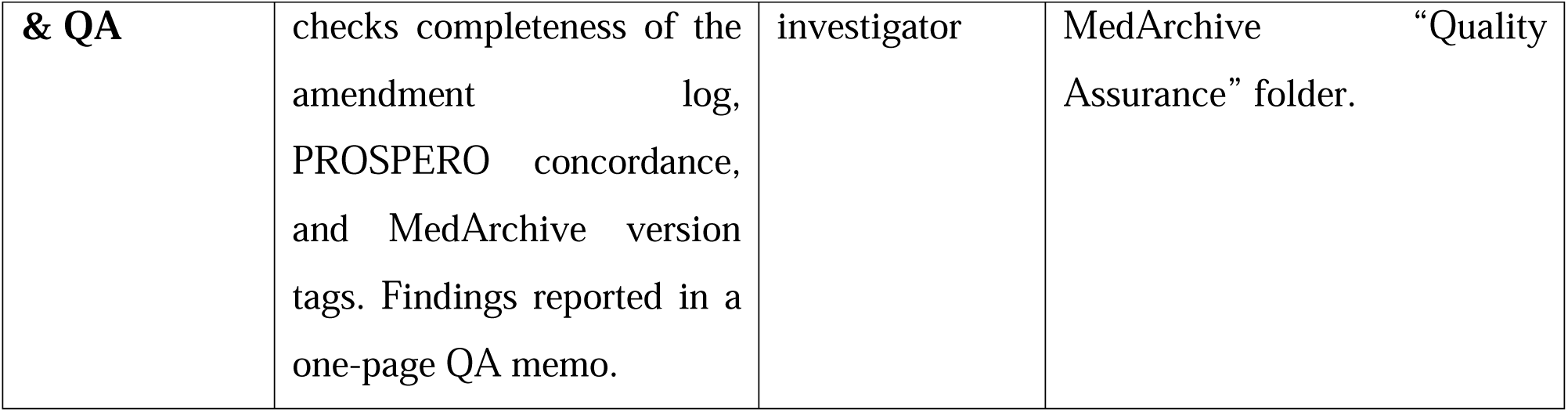

#### Contingencies

- If an amendment occurs *after* data extraction has begun, its potential to introduce bias will be assessed using a “decision impact” column in the log and, if substantive, addressed in the *Limitations* section of the final manuscript.
- Emergency amendments (e.g., database deprecation) may bypass the 72-h comment window but must be ratified retrospectively within one week.

